# Validation of a new approach for distinguishing anesthetized from awake state in patients using Directed Transfer Function applied to raw EEG

**DOI:** 10.1101/2020.06.18.20131508

**Authors:** Bjørn E. Juel, Luis Romundstad, Johan F. Storm, Pål G. Larsson

**Affiliations:** Brain Signaling Group, Institute of Basic Medical Science, University of Oslo, Norway; Department of Anesthesiology, Rikshospitalet, Oslo University Hospital, Norway; Department of Neurosurgery, Rikshospitalet, Oslo University Hospital, Norway

## Abstract

**Aim:** In a previous study, we found that the state of wakefulness in patients undergoing general anesthesia with propofol can effectively be monitored with high temporal resolution using an automatic measure of connectivity based on the Directed Transfer Function (DTF) calculated from short segments of electroencephalography (EEG) time-series. The study described here was designed to test whether the same measure can be used to monitor the state of the patients also during sevoflurane anesthesia.

**Methods:** Twenty-five channel EEG recordings were collected from 8 patients undergoing surgical anesthesia with sevoflurane. The EEG data were segmented into one second epochs and labeled as awake or anesthetized in accordance with the clinician’s judgement, and the sensor space directed connectivity was quantified for every epoch using the DTF. The resulting DTF derived connectivity parameters were compared to corresponding parameters from our previous study using permutation statistics. A data driven classification algorithm was then employed to objectively classify the individual 1-second epochs as coming from awake or anesthetized state, using a leave-one-out cross-validation approach. The classifications were made for every epoch using the median DTF parameters across the five preceding 1-second EEG epochs.

**Results:** The DTF derived connectivity parameters showed a significant difference between the awake and sevoflurane-induced general anesthesia at the group level (p<0.05). In contrast, the DTF parameters were not significantly different when comparing sevoflurane and propofol data neither in the awake nor in anesthetized state (p>0.05 for both comparisons). The classification algorithm reached a maximum accuracy of 96.8% (SE=0.63%). Optimizing the algorithm for simultaneously having high sensitivity and specificity in classification reduced the accuracy to 95.1% (SE=0.96%), with sensitivity of 98.4% (SE=0.80%) and specificity of 94.8% (SE=0.10%).

**Conclusion:** These findings indicate that the DTF changes in a similar manner when humans undergo general anesthesia caused by two distinct anesthetic agents with different molecular mechanisms of action. This seems to support the idea that brain connectivity is related to the level of consciousness in humans, although further studies are needed to clarify whether our results may be contaminated by confounding factors.

## Introduction

Objective quantification of how EEG signals change in relation to subjects’ states of consciousness has a long history (Loomis, Harvey, and Hobart 1937; Purdon et al. 2013; Brazier and Finesinger 1945). Recently, measures quantifying properties such as complexity (Casali et al. 2013), functional and effective connectivity (Lee et al. 2013), and information content (Schartner et al. 2015) in signals recorded with electroencephalogram (EEG) have been used successfully for objectively distinguishing between conscious and apparently unconscious states in humans. Generally, an apparent loss of consciousness is related to changes in such EEG signal properties or a combination of them (Mashour and Hudetz 2018), but capturing the relevant changes in a way that make the measures useful for bedside monitoring of the level of consciousness in patients is not straightforward.

Recently, we published results indicating that the Directed Transfer Function (DTF) – a measure that can be used to quantify sensor space directed connectivity from spontaneous EEG recordings – changed abruptly as patients undergoing surgical propofol anesthesia apparently lost and regained consciousness (Juel et al. 2018). The observed changes could be used to classify the state of individual patients as awake or anesthetized with 98% accuracy with a temporal resolution of 1 second. Importantly, these results were obtained using raw, spontaneous EEG data recorded in a normal clinical setting (general anesthesia for surgery), and without requiring any data cleaning, indicating that the DTF may be useful for developing objective, real-time monitors of patients undergoing anesthesia.

The previous study was done on a population of patients undergoing a single anesthetic protocol during surgery - propofol anesthesia, with the analgesic remifentanil. Thus, it is conceivable that the differences between the awake and anesthetized state observed in that study merely reflected changes related to the specific anesthetic agent rather than the changes related to general anesthesia, or loss of consciousness, in general. If so, the DTF-based method may not be fit to distinguish between states of consciousness more generally. However, qualitatively similar findings have been reported when using DTF to assess brain connectivity in groups of patients suffering from disorders of consciousness (Höller et al. 2014), and healthy individuals falling asleep (Kamiński, Blinowska, and Szelenberger 1997; Gennaro et al. 2004; Bertini et al. 2009). Taken together, this suggests that the DTF calculated from EEG may consistently change between conscious and unconscious states, regardless of how the change in state of consciousness came about.

Here, we report from a follow-up study, designed to test whether the DTF-based approach presented in our previous study can also be used to distinguish between awake and anesthetized states in patients undergoing general anesthesia caused by sevoflurane. Thus, we test whether the changes observed in the DTF derived connectivity parameters in the propofol study, were also apparent for patients undergoing sevoflurane anesthesia, and whether the changes could once again be used to successfully classify the state of the patients in accordance with the clinician’s judgement of their state of wakefulness.

## Methods

### Study design

This was a single-center observational study designed to investigate how the volatile anesthetic sevoflurane affects particular DTF derived connectivity parameters calculated from spontaneous EEG signals. The EEG data were collected from patients undergoing surgical sevoflurane anesthesia with fentanyl. The data were collected between September 2016 and February 2017 in experiments performed at the Oslo University Hospital, Rikshospitalet. Patients were recruited by the surgeon in charge of all included surgeries, the same surgeon as in our previous study (Juel et al. 2018). The study was approved by the Regional Committee for Research Ethics (case number 2012/2015), and all patients included in the study signed a written consent form after oral and written information.

### Inclusion and exclusion criteria

As in our previous study, the patients included were (1) American Society of Anesthesia I – III patients (ASA Physical Status Classification System. American Society of Anesthesiologists; https://www.asahq.org/resources/clinical-information/asa-physical-status-classification-system) (2) between 18 and 60 years old, and (3) seen as otherwise healthy based on a complete health examination. Patients were excluded if they had known hypersensitivity to sevoflurane or fentanyl, any history of, or family members with, malignant hyperthermia, soy oil or egg allergy, liver or renal disease affecting drug pharmacodynamics, heart or lung disease causing physical limitations (unable to climb two stairs without rest), body mass index greater than 30 kg/m^2^, any impaired general health condition from abuse of drugs and alcohol, organ damage, or neurological or psychiatric disease.

### Anesthetic management

The patients fasted for at least 6 hours before anesthesia. Their premedication consisted of oral paracetamol (Paracet®, Weifa, Oslo, Norway) 1.5 g, midazolam (Dormicum®, Basel, Switzerland) 3.75 – 7.5 mg for mild sedation, and oxycodone sustained release tablet (opioid analgesic; OxyContin®, Dublin, Ireland) 10 mg. Premedication was given 45 minutes before anesthesia. Before induction of anesthesia, an infusion with Ringer Acetat was started to compensate for any hypotension caused by anesthesia-induced vasodilatation and cardiodepression during induction. During anesthesia, the patients were monitored with pulse-oximetry (SpO2), and measurements of end tidal carbon dioxide (ETCO2), end tidal sevoflurane concentration (ETsevo), electrocardiography (ECG), and oscillometric noninvasive blood pressure (BP) every 5 minute. Anesthesia was induced with sevoflurane gas delivered via a vaporizer coupled to a semi-open breathing system and led to the patient through a tight face mask. The drugs used for anesthesia were sevoflurane, a non-pungent, non-irritable, ultra-short acting halogenated volatile general anesthetic (Sevofluran®,Baxter Medical AB Kista Sweden) and fentanyl 50 µg/ml, a potent, short-acting synthetic opioid analgesic (Fentanyl®, Hameln Pharmaceuticals Hameln Germany). After pre-oxygenation with 100 % oxygen 10 l/min for 3 minutes with spontaneous breathing in a tight face mask, fentanyl was given intravenously, and the sevoflurane vaporizer was set at maximum concentration of 8%. As the patient’s wakefulness and respiratory drive declined, the anesthesiologist started carefully to assist the ventilation with 8% sevoflurane in 100% oxygen (10 ml/min). When loss of eyelash reflex was observed, and the EEG had changed character from dominant alpha and low beta band activity to strong delta and theta/alpha activity in the frontal electrodes, the patient was intubated with an endotracheal tube. As soon as correct placement of the tube was verified, mechanical ventilation with 5% sevoflurane in medical air with 40% oxygen in nitrogen was started and the fresh gas flow was reduced from 10 to 2 l/min. We then varied the sevoflurane concentration between 5% and 3 % aiming for an end tidal sevoflurane concentration between 3% and 2% and 1 – 1.5 minimum alveolar concentration (MAC). Nitric oxide was not used. All the patients received local anesthetic infiltration with 5 ml 5% bupivacaine in the area of the skin incision.

### Assessment of consciousness

The patients’ state of wakefulness was assessed clinically by the anesthesiologist throughout the surgical procedure using standard anesthetic tools and practices. During the anesthesia induction phase, the Modified Observer’s Assessment of Alertness/Sedation Scale (MOAAS) (Chernik et al. 1990) was used to measure the patient’s state of wakefulness until loss of verbal contact and loss of response was reached. The MOAAS assessment was employed by the anesthesiologist maintaining verbal communication with the patient, and the patients were considered anesthetized and unconscious when they did no longer respond to their name being called (MOAAS level 2). At this point, the MOAAS assessment was discontinued, until the patient was about to wake up again. Throughout maintenance, the state of the patient was monitored clinically (by observing the heart rate, blood pressure, sweating, tear production, eye and eyelid reflexes, pupil size and symmetry, and any limb movements) with standard clinical equipment to ensure the conditions were suitable for all stages of surgery. Furthermore, the raw EEG, especially the recordings from the frontal electrodes, were observed providing information regarding the depth of anesthesia (Purdon et al. 2013). Time points for initiation and discontinuation of sevoflurane administration, loss of consciousness (LOC, i.e. corresponding to loss of verbal contact and behavioral response), and return of verbal communication (ROC) were recorded immediately by the electrophysiologist monitoring the EEG.

### EEG methods

EEG was recorded for the duration of the clinical procedure, including segments before, during, and after anesthesia. In total, 25 passive electrodes were used, 19 of which were placed in accordance with the 10-20 system (no mastoid electrodes), with six additional electrodes positioned to capture lower lateral activity (F9, F10, T9, T10, P9, P10). CP1 was used as the recording reference. No re-referencing was performed during the analysis.

The processing steps applied to the data closely resembled the analysis pipeline described in our previous paper (Juel et al. 2018). Each patient’s EEG data was read into Matlab using BioSig as implemented in EEGLAB (Delorme and Makeig 2004), and cut into non-overlapping 1-second epochs. The epochs from before LOC and after ROC were labeled as coming from the awake state, while the epochs between the markers for LOC and ROC were labeled as coming from the anesthetized state. Before further analysis, the data for each patient were automatically scanned for artefactual epochs and channels using a simple in-house algorithm based on the statistics of the patient’s own EEG signal. An epoch was marked as artefactual if it had large (deviating by more than 3 standard deviations from the median calculated from the patient’s own typical epochs signal) peak-to-peak amplitude, high variance, or transient currents (sudden changes in voltage) within the epoch. Similarly, a channel was marked as artefactual if its peak-to-peak amplitude, variance, or transient currents were different (again, deviating by more than 3 standard deviations from the median) when compared to other channels across epochs in the same patient’s EEG signal. The artefacts were not removed for the analyses but were labeled and used to indicate likely artefacts in figures (e.g. white patches in figures 4 and 6).

For all epochs, the relevant DTF variables were calculated (see (Juel et al. 2018) and (E. M. Schumacher, Stiris, and Larsson 2015)). DTF (for details, see: (Eichler 2006; M. J. Kaminski and Blinowska 1991)) was calculated for each 1 second epoch independently in the theta frequency range (4-8Hz), using the DTF function from the eConnectome toolbox (He et al. 2011). The median of the resulting matrices of DTF values was calculated across frequencies, yielding a typical strength of information flow between every pair of EEG electrodes in the theta band. The logarithm was taken to more clearly distinguish between small DTF values resulting in our main measure, referred to as LDTF. We also calculated the LDTF source strength (or information outflow) by taking the median across all outgoing connections from a given EEG channel. This was called mLDFT (or median information outflow) and represents the typical information each EEG channel apparently contains about the future activity in the other channels.

The LDTF values of each accepted epoch was then classified as coming from a segment recorded during the ‘awake’ or ‘anesthetized’ state using the classification algorithm presented in our previous work (Juel et al. 2018). The algorithm was based on a leave-one-out cross-validation scheme. Thus, for every patient, the LDTF matrix from each epoch was compared to the distributions of LDTF values pooled across all the other patients, for the awake and anesthetized state independently. The comparison yielded a value indicating the likelihood, L_state_, that the epoch LDTF values were drawn from each of the states (awake or anesthetized). The classification of the epoch, as either ‘anesthetized’ or ‘awake’, depends on the relationship between the likelihood values related to the two conditions. To quantify this relationship, we defined the ‘classification confidence’ (or ‘confidence of classifying an epoch as coming from the awake distribution’), C.

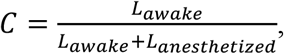

Here, L_awake_ is the likelihood that the LDTF values of the epoch were drawn from the (empirical) distribution of LDTF values from all other patients in the awake state. Similarly, L_anesthetized_ is the likelihood that the LDTF values of the epoch were drawn from the (empirical) distribution of LDTF values from all other patients in the anesthetized state.

We classified patient as awake or anesthetized in a given 1-second epoch depending on whether their C-value for that epoch was bigger or smaller than a preset threshold. To find the optimal value for this threshold, we varied the threshold between zero and one, and calculated the accuracy, sensitivity, and specificity of classification for each threshold value. The optimal value for C was defined as the threshold that produced the maximum sum of sensitivity and specificity across all patients.

### Statistics

The LDTF values were compared between states within each anesthetic protocol (awake vs anesthetized), as well as between types of anesthesia (sevoflurane vs propofol (from our previous study)). To do this, we calculated the mLDTF_awake_ and mLDTF_anesthesia_ for every patient (including patients from our previous study), and used permutation statistics method described in (Karniski, Blair, and Snider 1994). In brief, we quantified the difference between the mLDTF topographies (spatial maps of median information outflow from EEG channels) observed in the awake and anesthetized state, using the T-sum-squared statistic suggested by Karniski et al. Then, we made all possible permutations of groupings (switching the labels of mLDTF_awake_ and mLDTF_anesthesia_ for the patients in all possible ways) and quantified the difference between each of the groups using the same test statistic. If less than 5% of the permuted groups showed larger differences than the original grouping (e.g. awake vs anesthesia), the difference was considered significant. This was done for the sevoflurane patients as well as the propofol patients. We also did a permutation test with the data from propofol and sevoflurane pooled together, to compare the mLDTF topographies between states (awake and anesthetized) irrespective of anesthetic agent.

To investigate whether the changes observed under sevoflurane were comparable to changes observed under propofol, we ran two more permutation tests. Since the patients in the propofol study differ from the patients in the current study (thus, paired tests make no sense) the permutation tests were repeated 100 times, each time with a different ordering of the participants. In this way, we compared the mLDTF topographies in the propofol data (from previous study) with those in the sevoflurane data (from this study) within the awake and anesthetized conditions separately. This was done in order to test the hypothesis that the mLDTF topographies in comparable behavioral states were not different between the two experiments.

## Results

The final data material comprised 7 patients (3 female) with a median age of 48 years (range: 41-56 years). During the induction phase, the patients were intravenously given 350 µg (range: 250-450 µg) of the analgesic drug fentanyl, and the percentage of sevoflurane in the end-tidal volume was measured and adjusted (measured range: 2.6%-5.8%, mean 4.2%). After the patients became unresponsive no further doses of fentanyl were given. The median sevoflurane concentration during the maintenance phase of anesthesia was measured to 2.0% (range: 1.7%-2.4%) of end-tidal volume. Throughout the surgical procedure, physiological variables followed the expected development during anesthetic induction, maintenance, and emergence (Fig. 1).

**Figure 1:**
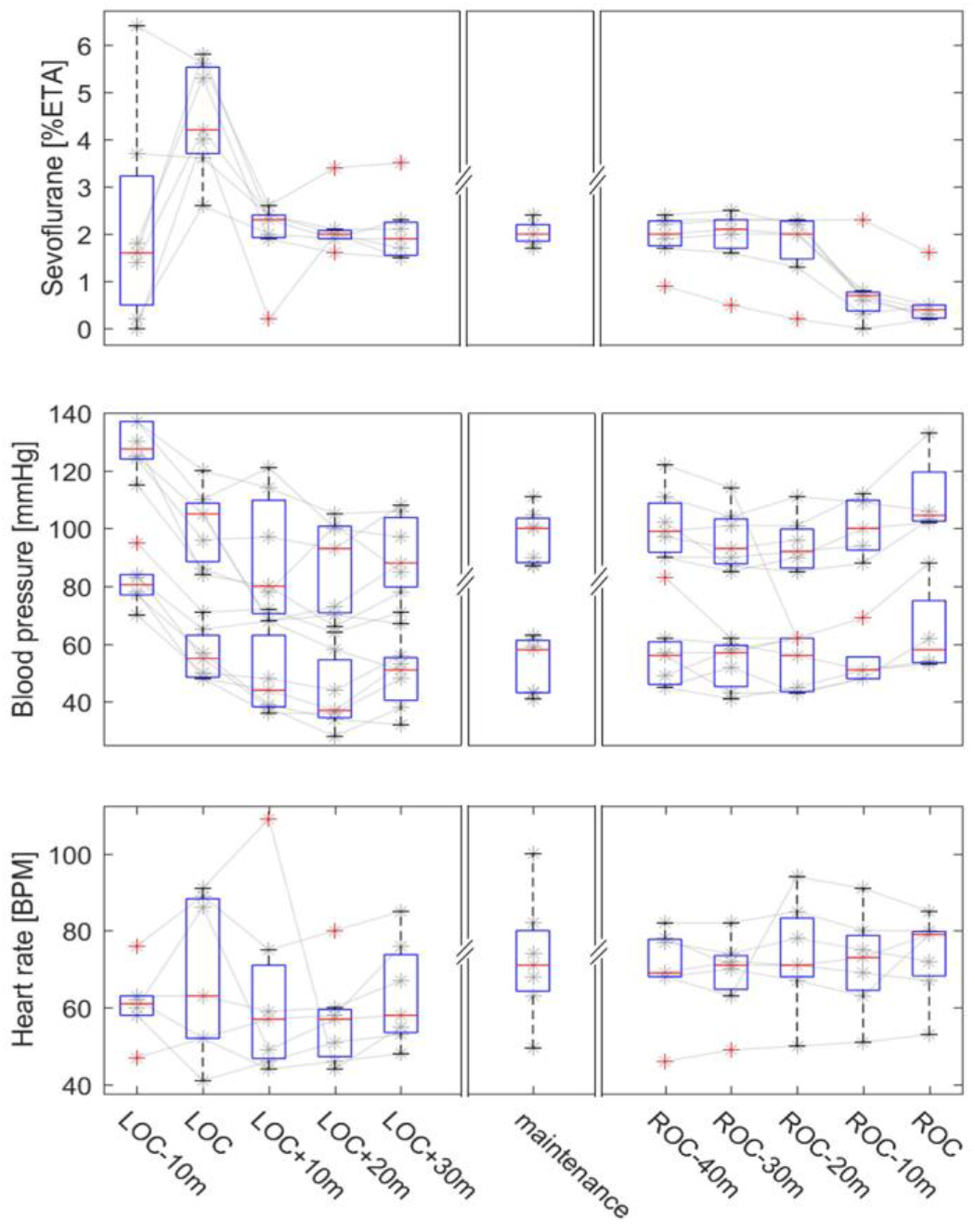
End-tidal sevoflurane concentration, blood pressure, and heart rate measurements during induction, maintenance, and re-emergence phases of surgical procedure. The panels show the relevant measures for each patient (gray lines and stars) with a box-and-whisker plot overlaid, summarizing the population statistics (red line: median, box: interquartile range, whiskers: minimum to maximum value, red crosses: outliers). Panel A shows the changes in percentage of sevoflurane in the end-tidal volume. Panel B shows systolic and diastolic blood pressure values. Panel C shows the heart rates. Each panel is subdivided into three parts, indicating the dynamics of each measurement during induction to (left) and reemergence from (right) anesthesia, and the mean values recorded throughout the maintenance phase (middle).

From each of the patients, continuous EEG was recorded throughout the clinical procedure (median length: 161 min, range: 115-247 min). Every recording contained segments from before (median length: 9 min, range: 7-16 min), during (median length: 148 min, range: 105-232 min), and after anesthesia (median length: 3 min, range: 2-5 min). In the automatic artefact scanning process, 13% (range: 2%-39%) of all epochs from the wakeful periods were marked as artefactual, while 6% (range: 4%-17%) of epochs were marked from the anesthesia period. In addition, a variable number of channels (median: 2, range: 0-4) channels were marked as artefactual.

The DTF analysis showed a qualitative difference between the ‘awake’ and ‘anesthetized’ states. The mLDTF topography was heterogeneous in the ‘awake’ state, but more homogenous in the ‘anesthetized’ state (Fig. 2; panel B). A relatively strong apparent source of information outflow could be seen located over the posterior midline channels, whereas this region was far less distinct in the plot for the ‘anesthetized’ state. This looks similar to the changes observed in our previous work with patients undergoing propofol anesthesia (Fig. 2; panel C).

**Figure 2:**
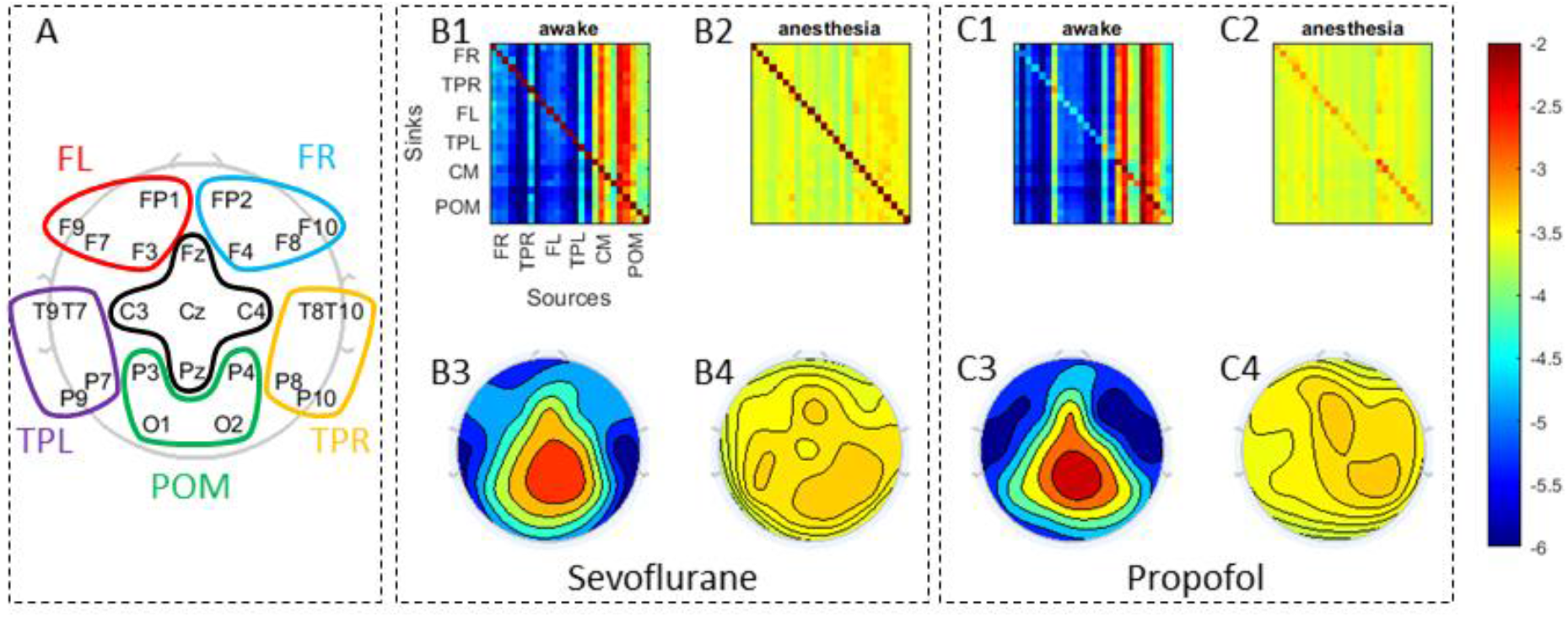
Summary figure showing population median DTF connectivity patterns. Panel A shows a topographical representation of the channel montage used during the EEG recordings. For clarity, sets of electrodes grouped together and marked by colors in order to simplify the displays in the other two panels (Frontal Right (FR) electrodes are marked in Blue: FP2, F4, F8, and F10; Temporoparietal Right (TPR) electrodes are marked in Orange: T8, T10, P8, and P10; Frontal Left (FL) electrodes are marked in Red: FP1, F3, F7, and F9; Temporoparietal Left (TPL) electrodes are marked in Yellow: T7, T9, P7, and P9; Central Medial (CM) electrodes are marked in Black: Fz, Cz, C4, C3, and Pz; and Posterior Occipital Medial (POM) marked in Green: Pz, P4, P3, O2, and O1). In panels B and C the population median DTF in the theta range is visually summarized for the sevoflurane (new data) and propofol (data from Juel et al. (2018)), respectively. Panels B1 and C1 show the full directed connectivity matrices in the awake state, while B2 and C2 show the same for the ‘anesthetized’ state. Each element in the matrix quantifies the median information flow from a source channel (x-axis) to a sink channel (y-axis). In the corresponding topographical plots (B3-B4; C3-C4), the distribution of information flow sources across the scalp are shown. B3 and C3 visualize how DTF-based information sources are distributed across the scalp in the awake state. B4 and C4 do the same for information sources in the anesthetized state. The color scale on the right indicates the relation between the colors used and the values of LDTF for all panels: red indicates strong, while blue indicates weak, information flow.

The mLDTF topographies from each individual, in both awake and anesthetized states, can be seen in Figure 3. Permutation tests indicate that the qualitative differences observed between conditions were statistically significant (p=0.016; 2^7=128 permutations) for the patients undergoing sevoflurane anesthesia. A significant change was also observed when comparing the awake with the anesthetized condition in the data from the patients undergoing propofol anesthesia (p=0.016; 2^7=128 permutations). This was also the case when pooling together the data from the two experiments, indicating that the differences observed here might be similar to those observed in our previous study (p=4.0*10^-4; 2^14=16384 permutations). In all tests, the natural grouping of patient labels (anesthesia vs awake) was the grouping with the largest overall difference between the groups of all possible permutations, indicating the strongest possible evidence for differences between groups given the number of samples. Furthermore, when comparing within conditions, between experiments (propofol anesthetized vs sevoflurane anesthetized and propofol awake vs sevoflurane awake), the test yielded non-significant results (p>0.05 for all 100 runs; 2^7=128 permutations). Thus, there was little or no evidence for differences between mLDTF values in the patient groups from the two studies, in either the anesthetized or the awake state, and we cannot reject the hypothesis that the mLDTF topographies in comparable behavioral states were not different between the two experiments.

**Figure 3:**
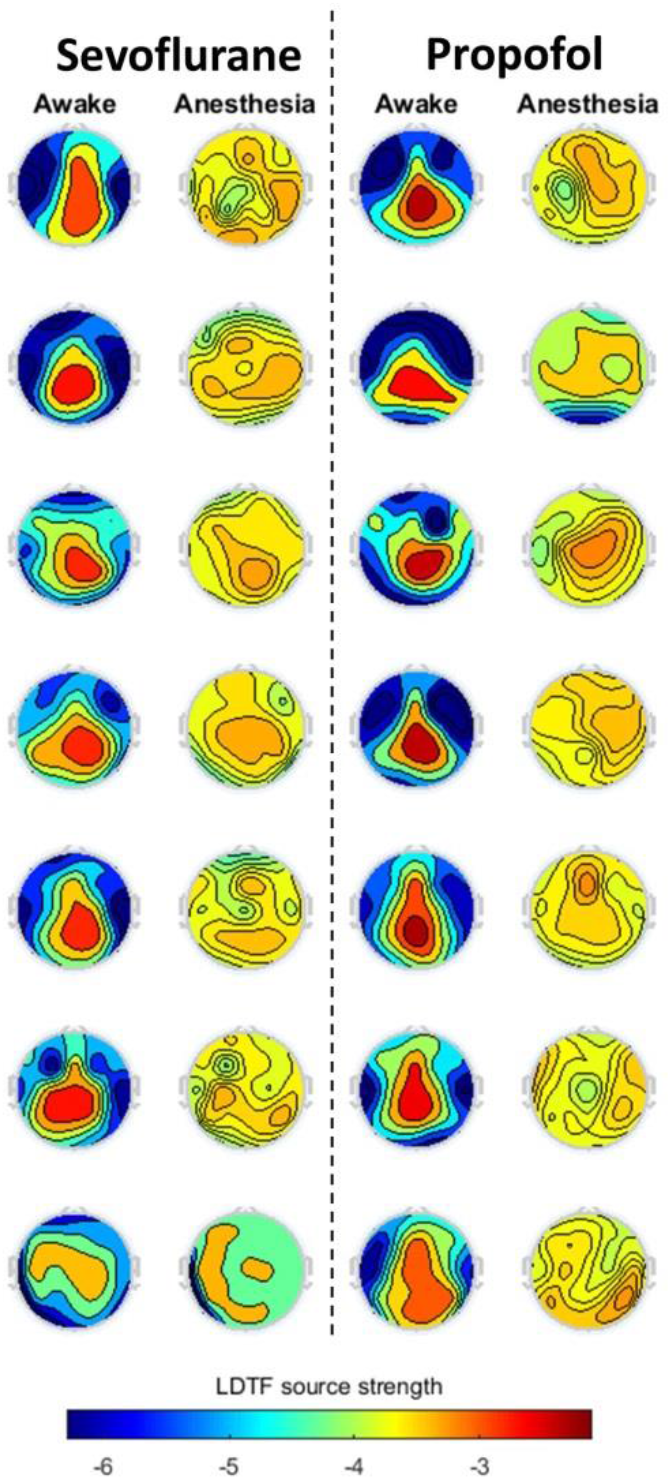
Topographical maps of DTF-based median outgoing connectivity strengths in the awake and anesthethized state. The topographical map of mLDTF from the sevoflurane (left, theta range mLDTF) and propofol (right, alpha range mLDTF; data from Juel et al. (2018)) studies are shown. Each panel shows the pooled mLDTF map for a given patient (row) in a given state (column). The color scale on the bottom indicates the relation between the colors used and the values of LDTF for all panels: red indicates strong, while blue indicates weak, DTF-based information flow.

The mLDTF patterns remained relatively stable over time within states, but of the patterns transitioned abruptly when the patients’ state of wakefulness changed (Fig. 4). This was the case both when patients transitioned from wakefulness to anesthesia (Fig. 4, left column) as well as when the patient regained consciousness after anesthesia (Fig. 4, right column). However, there was no distinct change in the mLDTF pattern around the time when the anesthesia delivery was stopped (Fig. 4, middle column), Importantly, the abrupt change in mLDTF pattern was clearly reflected in the algorithm’s ‘classification confidence’ (black lines in Fig. 4), which appears to transition between two distinct states right around the transitions between wakefulness and anesthesia.

**Figure 4:**
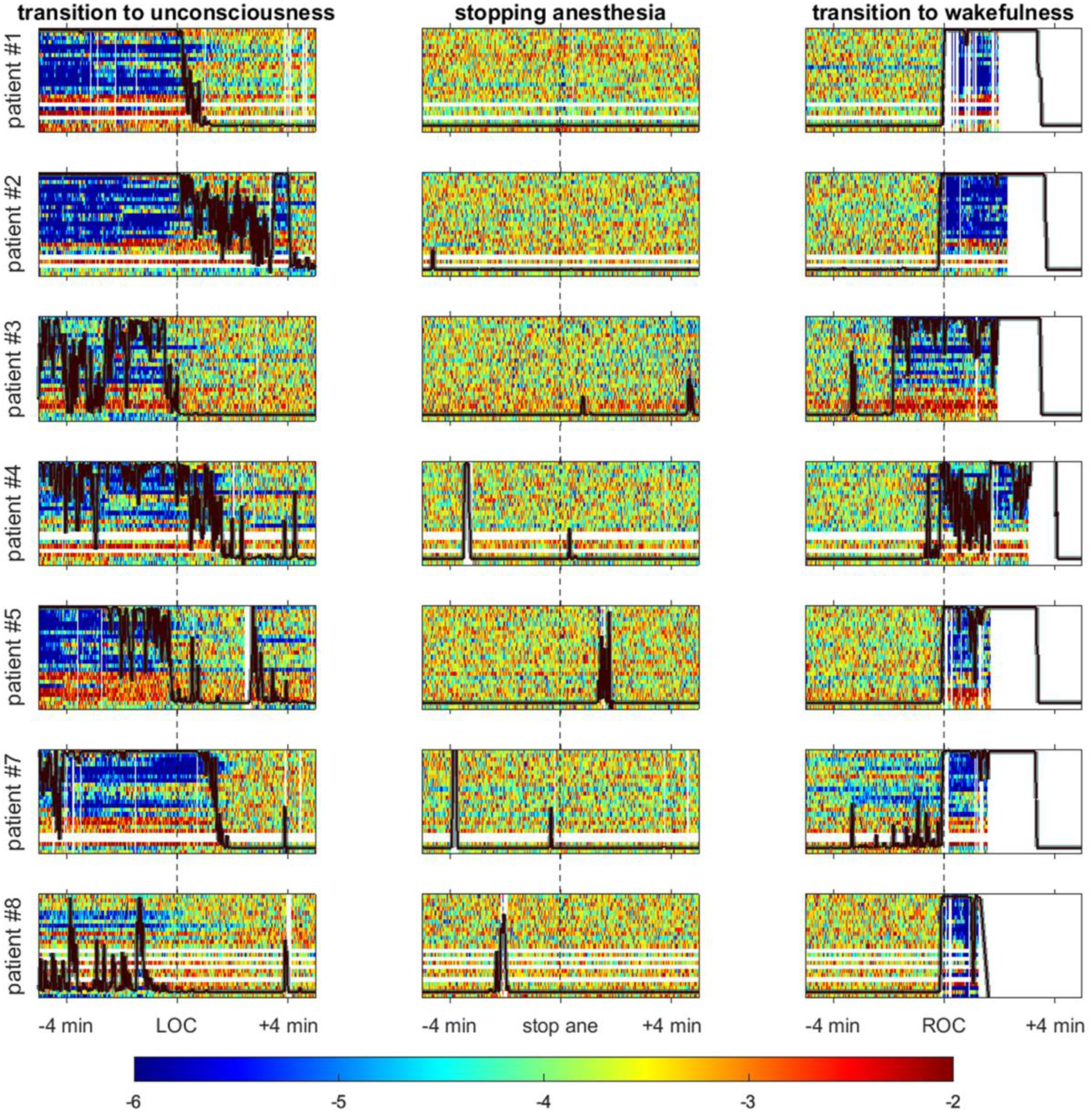
Visualizing the LDTF source strengths, and the classification confidence, near main events in the anesthetic management. Each row in this figure contains three plots useful for describing the quality of the classification algorithm. The figures in each row shows how the information source strengths change for each patient around three critical points in the anesthetic management – loss of verbal communication (LOC, left), stopping the anesthetic administration (stop ane, centre), and the time of return of verbal communication (ROC, right). Each plot is time locked to the time-point noted by the clinical staff, and shows the development of the mLDTF source strengths from five minutes before, to five minutes after the event. White vertical lines indicate an artefactual epoch, and white horizontal lines indicate an artefactual channel. The colors in all plots relate to the strength of the information source as indicated by the color bar.

From the LDTF values, the algorithm’s confidence (C) in classifying a patient as awake in a given one-second epoch was calculated to form a basis for classification. Varying the threshold value required for classifying a patient as awake resulted in a maximum overall accuracy of 96.8% (SE=0.63%) for threshold values of 0.1<C<0.17 (Figure 5). The classification accuracy remained high for a broad range of threshold values, only dropping below 96% for C<0.01 and C>0.9. However, when we optimized for maximized sum of sensitivity and specificity, the range of valid threshold values shrank significantly, and moved towards lower values of C. Specifically, the optimal threshold value was C=0.001, yielding an accuracy of 95.1% (SE=0.001), with sensitivity of 98.4% (SE=0.80%) and specificity of 94.8% (SE=0.10%). The results of the classification using the optimal threshold value is shown in Figure 6 to give an impression of the temporal stability of the mLDTF patterns and the accuracy of the classification for every patient.

**Fig 5:**
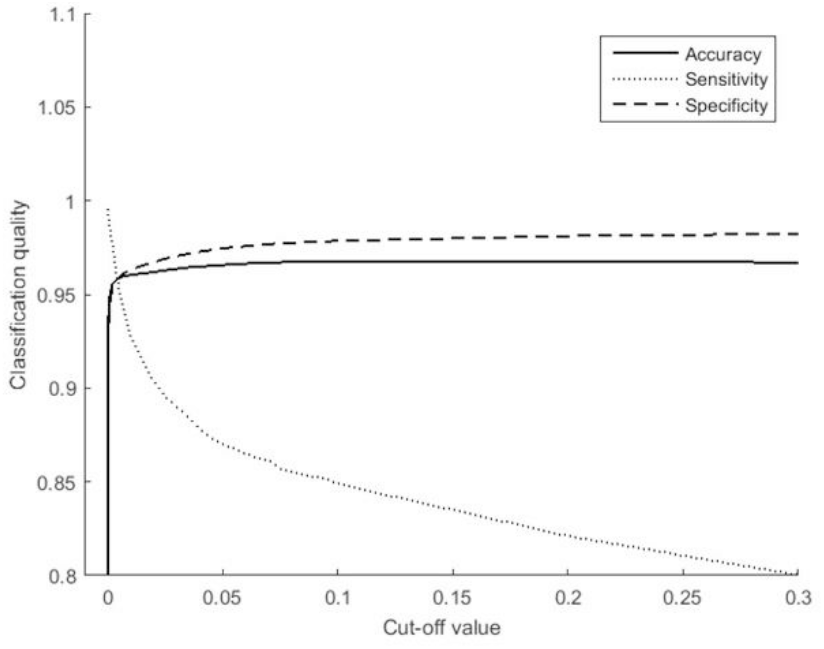
The accuracy, sensitivity, and specificity of classification is plotted against cut-off values of the parameter C, above which the algorithm would classify the patient as conscious. The accuracy of the classification is relatively stable for a broad range of thresholds, but the sensitivity rapidly drops of as the threshold increases.

**Fig 6:**
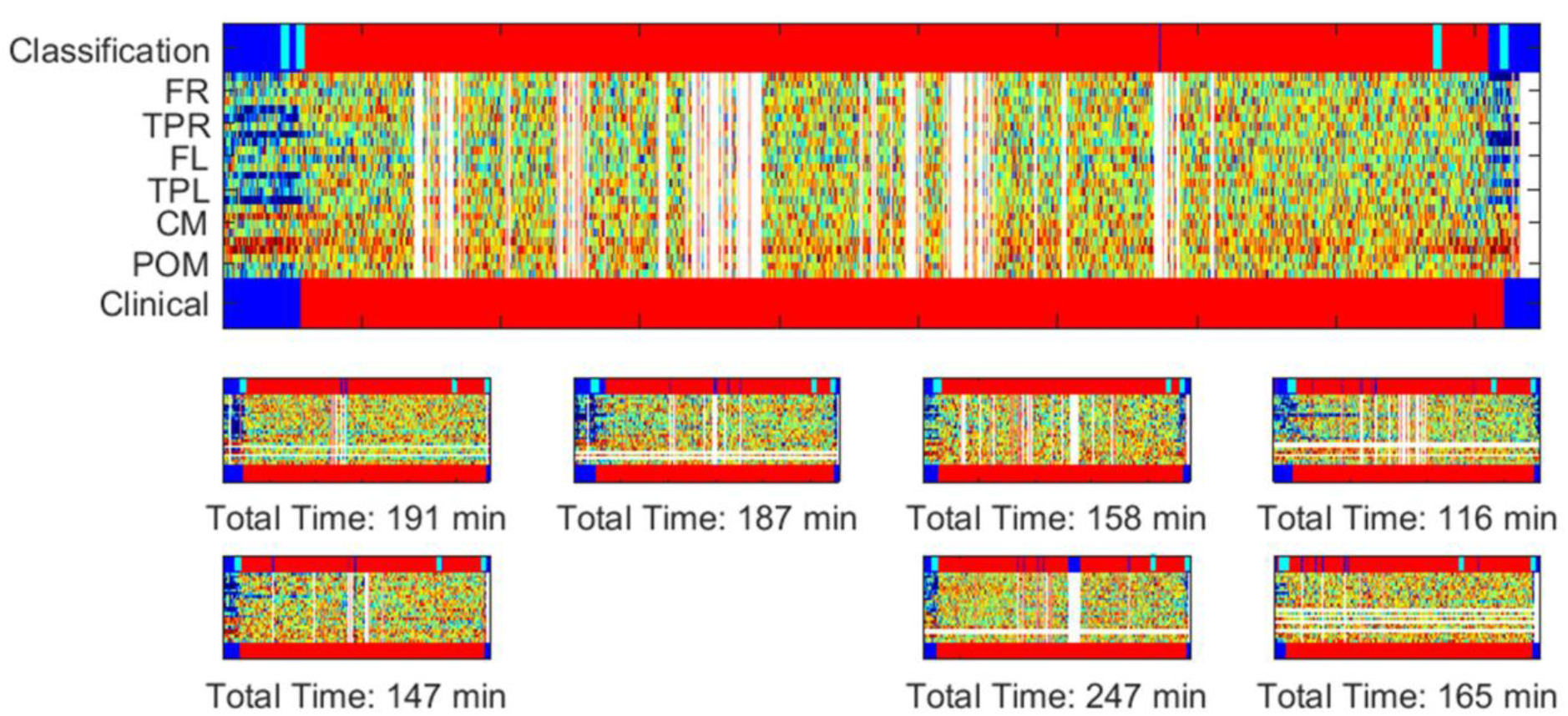
Visualization of classification results for each patient. The time courses of DTF information source strengths for all patients are shown, together with their corresponding clinical judgement, and algorithmic classification, of their conscious state. The middle region of each panel, containing the information source values (LDTF) for every channel, follows the color scheme indicated in the color bar. In addition, epochs and channels marked as artefactual by the automatic data cleaning algorithm are marked with white columns and rows respectively. The bottom bar indicates the states of the patient reported by the clinical staff (blue: awake, red: anesthetized). The top bar represents the corresponding conscious state of the patient as classified by the algorithm (using the optimal threshold for classifying as awake, C>0.001). In each panel, turquoise lines indicate the four main events in the anesthetic management: start of anesthesia administration, loss of verbal communication, stopping the anesthesia administration, and return of verbal communication. Panel A shows the same information as panel D (patient 3), just enlarged to give a better impression of the dynamics, as well as details in the classification.

## Discussion

This study shows that our DTF-based measure yields similar results when applied to EEG recordings obtained from patients undergoing general anesthesia with sevoflurane and with propofol. Like we found with propofol anesthesia in our previous paper (Juel et al. 2018), the source strength topographies went from having relatively stronger apparent sources of information outflow in the posterior region of the head in the ‘awake’ state, to becoming far more homogeneously distributed in the ‘anesthetized’ state during sevoflurane anesthesia (Fig. 3). The differences between conditions (awake vs. anesthesia) were significant for both anesthetics (propofol and sevoflurane), and there was no significant difference between the changes observed with the two anesthetics. Taken together, this indicates a common change being captured by the LDTF when comparing the awake and anesthetized conditions, using different anesthetics, under slightly different conditions, and using slightly different analysis. Finally, the changes in DTF-based parameters could be successfully used to classify the patients’ states as awake or anesthetized in accordance with the clinician’s judgement with a 1-second temporal resolution.

Even though these results are similar to what was found in our previous study, there were some differences in the analysis that should be mentioned. In addition to adding an automatic artefact-scan (only used for visualization/reporting), we changed two aspects in the analysis: in the present study we (1) focused on the theta rather than alpha frequency band, and (2) optimized the classification using the parameter C – the ‘confidence of classifying an epoch as coming from the awake distribution’.

We changed the frequency band used for DTF analysis from the alpha band, which we used in our previous study (Juel et al. 2018) to the theta band, to reduce potential confounds caused by changes in the alpha band power related to closing of the eyes (patients had their eyes open in the awake state but closed in the anesthetized state). The patterns of DTF derived connectivity observed in the two bands were relatively similar in our previous analyses leading us to believe that it would be possible to successfully use the theta band for the purpose of distinguishing between the states. Indeed, this change seems to yield a small benefit in reducing one potential confound, without causing any clear disadvantages or detrimental effects to the classification. Similarly, the choice of optimizing the threshold parameter, C, seemed to improve the analysis compared to use the more naïve approach used for classification from our previous paper. Interestingly, this optimization had quite a strong effect on the quality of classification in the current paper. Even though the accuracy of classification would have been similar using the naïve approach, the sensitivity was improved when using the optimized threshold (see Figure 5). In fact, the sensitivity would have been worse than in our previous paper if we had not used a different threshold (only ∼70% of the epochs from awake patients would have been correctly classified, in contrast to the 95% in our previous paper). However, using the optimized threshold, C=0.001, instead yielded classification results that were slightly better than in our previous study (Juel et al. 2018).

Another difference from the previous study was that the patients were asked about whether they experienced anything during the anesthesia and surgical procedure, to investigate whether any of them would have recollection of waking up, dreaming, or otherwise being conscious. However, to avoid inducing traumatic memories, the depth of the questioning was kept to a minimum. None of the patients reported memories of waking up during the surgery, but two patients (#1 and #8) reported having had simple dreams during the anesthesia. However, their timelines of typical outgoing information flow (mLDTF) did not show any sign of awake-like patterns during the anesthesia. There might be several reasons for this, but we do not have sufficient data to make strong conclusions here. For example, our measure may be insensitive to the dream state, the dreams may have occurred during emergence or have been confabulated, or the patients may have had brief periods of waking up that were later interpreted as dreaming or forgotten. Determining the real causes of this sort of observations requires a different type of protocol, better suited for experiments outside of clinical surgery setting.

Of course, the most profound difference between this study and the previous was the fact that the patients underwent a different type of anesthesia (sevoflurane rather than propofol). This fact was used to investigate whether the DTF-method can successfully classify the state of wakefulness in patients using an anesthetic with assumed distinct mechanism of action (Alkire, Hudetz, and Tononi 2008; Rudolph and Antkowiak 2004), but a comparable endpoint for the patient: unresponsiveness and apparent unconsciousness due to the general anesthetic (Hudetz and Mashour 2016). The two anesthetics, propofol and sevoflurane, share some mechanisms of action: they both potentiate GABA_A_ and glycine receptors, and inhibit voltage gated potassium channels and acetylcholine receptors (Rudolph and Antkowiak 2004; Alkire, Hudetz, and Tononi 2008). However, they also differ in certain respects. Most notably, sevoflurane potentiates two-pore potassium channels and inhibits serotonin receptors, while propofol potentiates kainate receptors. Furthermore, sevoflurane has been reported to have a stronger inhibiting effect on AMPA and NMDA receptors. In short, the two anesthetics have complex and different interactions with ion channels and receptors affecting several neuronal populations, thus profoundly altering the neuronal activity patterns in the brain (Antkowiak 1999).

These alterations in neuronal activity are reflected in changes in EEG patterns. For example, as the molecular targets and effects of sevoflurane and propofol both partly overlap and partly differ, there are both similarities and differences also between their effects on large-scale measures of brain function (Gugino et al. 2001). For example, both sevoflurane and propofol have been reported to induce coherent frontal alpha oscillations and slow oscillations in EEG of humans (Akeju et al. 2014), but sevoflurane has also be shown to be unassociated with the typical anteriorization of alpha rhythms (Blain-Moraes et al. 2015). Sevoflurane anesthesia has also been seen to increase the coherence in the theta frequency range (4-7Hz), relative to comparable levels of propofol anesthesia (Akeju et al. 2014). In addition, propofol and sevoflurane have been shown to differentially suppress the relative glucose metabolic rate in several bran regions (Jeong et al. 2006). Furthermore, it is well known that anesthetics affect somatosensory evoked EEG potentials in humans and animals (Banoub et al. 2003), but sevoflurane affects these potentials more strongly, in a dose-dependent fashion, than comparable doses of propofol (Boisseau et al. 2002, Arena et al. 2016). These findings indicate that certain large-scale properties of the EEG do indeed change differentially in response to the two anesthetics. The fact that both the molecular, cellular, and large-scale brain effects of the two anesthetics differ in so many ways, increases the value of testing our method with both compounds, and enhances the significance of the remarkable similarity in their effects on DTF. Thus, since it is not obvious that EEG-derived measures such as the DTF would yield so similar results in patients undergoing anesthesia with as mechanistically different agents as propofol and sevoflurane, our results suggest that they have some substantial large-scale effects in common that are captured by the DTF derived measure.

In recent years, results from a broad range of studies seem to be converging on the “conclusion that a common neural correlate of anesthetic-induced unresponsiveness is a consistent depression or functional disconnection of lateral frontoparietal networks, which are thought to be critical for consciousness of the environment” (Hudetz and Mashour 2016). DTF is one of several measures that can be used to quantify aspects of large-scale connectivity between time series such as EEG signals (Blinowska 2011). And, in addition to our own previous study (Juel et al. 2018), at least four studies have previously investigated how DTF-based connectivity measures are affected by changing states of consciousness (Kamiński, Blinowska, and Szelenberger 1997; Bertini et al. 2009; Höller et al. 2014; Gennaro et al. 2004). Each one of them reported changes in apparent brain connectivity related to distinct states of consciousness. Bertini et al. reported that the DTF-based connectivity changed between wakefulness and stage 2 sleep (Bertini et al. 2009). Specifically, they report that the “inter-hemispheric directional flows varied as a function of the state of consciousness”. The observed change was particularly clear in the alpha frequency band between the parietal electrodes P3 and P4. In our study, the largest changes in connectivity are also apparent in the posterior electrodes. In another study, Gennaro et al. (Gennaro et al. 2004) showed that the DTF-based alpha range connectivity between posterior and frontal electrodes was inverted at sleep onset. Thus, in the pre-sleep period, connectivity was stronger from parietal to frontal (Pz to Fz) electrodes than the other way around, while this asymmetry was inverted at sleep onset. Similarly, Kaminski et al. (Kamiński, Blinowska, and Szelenberger 1997) also observed changes in the DTF-based connectivity at the onset of sleep. Specifically, they reported a “diminishing role of the posterior sources and an increasing effect of the anterior areas”, which is reminiscent of the changes we observed at the onset of general anesthesia with propofol and sevoflurane (Figure 3). Again, these reports emphasize the loss of back-to-front connectivity related to the apparent loss of consciousness. Using a different approach, Höller et al. (2014) found that differences in DTF were among the most effective measures for distinguishing between unresponsive and minimally conscious states in patients with disorders of consciousness (DOC). Taken together, these reports provide evidence that changes in DTF may capture properties related to loss of consciousness in general, not just specific features of general anesthesia caused by sevoflurane (this study) or propofol (Juel et al. 2018).

In addition to the DTF, several other measures of connectivity have been suggested as a relevant markers for changes in the state or level of consciousness, including measures of transfer entropy (Ku et al. 2011; Lee et al. 2013), directed coherence (Lioi et al. 2017; Maksimow et al. 2014), and Granger causality measures (Barrett et al. 2012; Seth, Barrett, and Barnett 2011). Specifically, measures of the connectivity between frontal and parietal regions were found to differ between conscious and unconscious states, both for humans undergoing various forms of anesthesia (Ku et al. 2011; Lee et al. 2013) or falling asleep (Lioi et al. 2017), and for patients suffering from DOC (Boly et al. 2012). For example, in a study using directed coherence – a measure closely related to DTF – the directionality of frontal-parietal functional connectivity covaried with NREM sleep stages and wakefulness (Lioi et al. 2017). However, approaches such as these (ours included), trying to quantify network properties based on passive observation of brain activity, are unlikely to justify strong conclusions about the underlying brain mechanisms (Massimini et al. 2009). Perturbational approaches, however, have indicated that a balanced interconnectivity between distant brain regions is likely to be crucial for maintaining a normal capacity for consciousness (Massimini et al. 2005; Ferrarelli et al. 2010; Rosanova et al. 2012; Casali et al. 2013; Casarotto et al. 2016). Thus, it is plausible that reports from observational studies of altered connectivity related to loss of consciousness also often reflect relevant underlying changes.

The fact that filtering, rejection of artefacts, and other data cleaning techniques were deliberately avoided in this study (in order to simulate a setting relevant for real-time monitoring of brain states in the clinic) may have further distorted the resulting connectivity matrices and yield faulty estimations of brain connectivity patterns. However, since the DTF is known to be robust to noise (Astolfi et al. 2007), and the median was used as a descriptive statistic for the mode of the distributions (Dukic et al. 2017), this type of problems should be minimized (Schumacher et al. 2011). Nevertheless, we remain agnostic regarding how the results reported here (regarding scalp level inferred connectivity) are related to the underlying changes in neural, effective connectivity within the brain. In fact, even with properly cleaned EEG data, the degree to which of sensor space estimates of connectivity are relevant for characterizing the underlying neural connectivity is disputed (Papadopoulou, Friston, and Marinazzo 2015; Brunner et al. 2016; M. Kaminski and Blinowska 2017).

Furthermore, it is possible that our DTF measure may be influenced by changes in muscle activity related to anesthesia, as has previously been shown for the bispectral (BIS) index which is one of several methods used for monitoring depth of anesthesia (e.g. Schuller et al. 2015; Zetterlund et al. 2016). Propofol is known to decrease muscle tone in the absence of neuromuscular blocking drugs (Haeseler et al. 2001), and it is uncertain to what extent such effects may contribute to the observed changes in DTF reported here and in our previous paper (Juel et al. 2018) as it may not be not possible with our methods to tease apart muscle relaxing effects from other effects anesthetics have on the brain and body. Similarly, sevoflurane is known to cause immobility (King and Rampil 1994), depress the excitability of motor neurons (Zhou, Mehta, and Leis 1997), and enhance the effect of neuromuscular blockers (Wulf, Kahl, and Ledowski 1998). However, the degree to which sevoflurane affects muscle tonus and EMG contamination of the EEG signal under the conditions used in our study remains to be determined. Thus, we cannot exclude the possibility that the apparent changes in brain connectivity observed here, may at least partly result from changes in EMG contamination in the two states, rather than reflecting consciousness related changes within the brain. This is an issue that is hard to control for without additional pharmacological interventions that were not feasible in the clinical setting our data were obtained from. We did, however, avoid neuromuscular blockers when possible to minimize anesthesia related changes in EMG (only one patient (#7) received an additional neuromuscular blocker (cisatrakurium, 14 mg i.v.) due to problems related to intubation). However, to investigate this issue more directly, we are now initiating a follow-up study in which we will test the effect of pure neuromuscular block in the absence of anesthetics on measures assumed to track the depth of anesthesia.

Another possible limitation is that our choice of DTF as a measure was made for reasons not entirely grounded in theories of consciousness. This makes it more likely that the observed changes in connectivity are related to changes in state more generally (e.g. going from normal wakefulness to general anesthesia) rather than reflecting changes in the brain that are specifically and causally important for consciousness. However, taken together, our findings that similar patterns of DTF changes occur for at least two different types of anesthesia, combined with the previous findings of changes in DTF related to distinct states of consciousness, provide convergent evidence in support of a strong relation between consciousness and measures of brain connectivity (e.g. Höller et al. 2014; Kamiński, Blinowska, and Szelenberger 1997; Gennaro et al. 2004; Bertini et al. 2009), although further studies are needed to clarify whether and how the observed changes in our DTF-based measure are related to changes in consciousness as such.

Finally, other efforts are ongoing to reproduce our previous findings (Juel et al., 2018) in different experimental settings and under different conditions in order to test the generalizability of the DTF-based measure as a marker of consciousness. However, these efforts have shown that reproduction is not always straight-forward, and that the particular topography of apparent information outflow may be sensitive to parameter choice in the analysis (e.g. filter types, sampling frequency, reference position, spatial filter functions) as well as the state of the person having their EEG measured. Thus, before drawing strong conclusions about the capacity of the method presented here and in our earlier publication to track conscious states in humans, further studies are needed.

## Conclusions

Following our previous study where a DTF-based approach was used to detect changes in apparent brain connectivity during general anesthesia with propofol, we performed a follow-up study to validate our method by testing whether similar changes occur during loss of consciousness caused by another anesthetic, sevoflurane, which has partly different mechanisms of action. We found that our DTF-derived connectivity parameters showed similar changes in patients undergoing sevoflurane anesthesia as were previously observed with propofol anesthesia. These changes could once again be used to successfully classify the patients’ state of anesthesia vs. wakefulness in accordance with the clinician’s judgement, with accuracies and sensitivities exceeding 96%, depending on the choice of cut-off value for our algorithm’s confidence in classification. These results indicate that certain changes in DTF caused by general anesthesia generalize across at least two different anesthetics with partially distinct mechanisms of action. This can be regarded as further evidence in favor of brain connectivity being related to the level of consciousness in humans, although our study does not yet exclude the possibility that effect on muscle activity (EMG) may contaminate our EEG results. Thus, further studies are required for better understanding how the observed alterations in DTF are related to changes in brain state and consciousness.

## Data Availability

The data used for this work are not available online, but may be made available for individuals upon request.

## Acknowledgement

We would like to thank Frode Kolstad for being central in setting up the study, selecting patients and performing all the surgeries. Furthermore, we would like to thank Sally Rose and Marianne Nævra for their contributions to the study in mounting and recording the EEG, and handling of the data. This study was supported in part by the European Union’s Horizon 2020 research and innovation programme under grant agreement 7202070 (Human Brain Project (HBP)) and the Norwegian Research Council (NRC: 262950/F20 and 214079/F20).

